# Hierarchical integration of multimodal clinical data to predict epilepsy surgery outcome

**DOI:** 10.64898/2026.05.05.26352481

**Authors:** John Thomas, Chifaou Abdallah, Thandar Aung, Pilar Bosque-Varela, Irena Doležalová, Prachi Parikh, Lara Wadi, Kassem Jaber, Zhengchen Cai, Alyssa Ho, Mathew K Moye, Erica Minato, Olivier Aron, Stephan Chabardes, Sophie Colnat-Coulbois, Jeffery Hall, Petr Klimes, Lorella Minotti, François Dubeau, Derek Southwell, David Carlson, Milan Brazdil, Jorge Gonzalez-Martinez, Philippe Kahane, Louis Maillard, Jean Gotman, Birgit Frauscher

**Affiliations:** Department of Electrical and Computer Engineering Technology, Rochester Institute of Technology, NY, USA; Department of Biomedical Engineering, Duke Pratt School of Engineering, Durham, NC, USA; Montreal Neurological Institute and Hospital, McGill University, Montréal, Québec, Canada; Department of Neurology, University of Pittsburgh Medical Center, Pittsburgh, PA, USA; Department of Neurology, Neurocritical Care and Neurorehabilitation, Christian Doppler University Hospital, Member of the European Reference Network EpiCARE, and Centre for Cognitive Neuroscience, Paracelsus Medical University, Salzburg, Austria; Neuroscience Institute, Christian Doppler University Hospital, Centre for Cognitive Neuroscience, Salzburg, Austria; Department of Neurology, Duke University Medical Center, Durham, NC, USA; Brno Epilepsy Center, First Department of Neurology, St. Anne’s University Hospital, Faculty of Medicine, Masaryk University, Brno, Czech Republic; Cumming School of Medicine, University of Calgary, Calgary, AB, Canada; Department of Neurology, University Hospital of Nancy, Lorraine University, F-54000 Nancy, France; IMOPA, Clinical research team in neurosciences, Lorraine University, CNRS, UMR, 7365 Vandoeuvre, France; CHU Grenoble-Alpes, Université Grenoble Alpes, Inserm, U1216, Grenoble Institute Neurosciences, Grenoble, France; Department of Neurosurgery, Duke University Medical Center, Durham, NC, USA; Department of Biostatistics and Bioinformatics, Duke University School of Medicine, Durham, North Carolina, USA; Department of Computer Science, Department of Civil and Environmental Engineering, Duke University, Durham, North Carolina, USA; Department of Neurosurgery, University of Pittsburgh Medical Center, Pittsburgh, PA, USA

**Author notes:** Corresponding author: Dr. Birgit Frauscher, Dr. John Thomas, Corresponding author’s address: Duke University Medical Center, Department of Neurology, 2424 Erwin Street, Ste 1001, Durham, NC, 27705, Corresponding author’s, Corresponding author’s e-mail address. listed in alphabetical order for equal contribution (Thandar Aung, Pilar Bosque-Varela, Irena Doležalová, Prachi Parikh, Lara Wadi).

## Abstract

**Background:** Integrating multimodal data into medical artificial intelligence (AI) tools and evaluating whether they outperform human experts remains a critical challenge. Epilepsy surgery offers a unique paradigm for this evaluation, as it provides an expert-independent measure (Engel score) of post-surgical outcome. Currently, evaluation for epilepsy surgery relies on the visual interpretation and human synthesis of multimodal data. While clinical evaluations are individualized and account for complex anatomical variability, integrating these diverse, high-dimensional modalities to generate a probability of surgical success remains challenging. Here, we leverage this objective outcome score to investigate the feasibility of a data-driven, phenotype-based model against the current clinical gold standard.

**Methods:** The evaluation was performed on an epilepsy-type controlled cohort of 57 patients from six tertiary epilepsy surgery centers who underwent resective/ablative surgery in the mesiotemporal lobe. Multimodal data, namely, patient demographics, semiology, invasive electrophysiology monitoring, and neuroimaging, were utilized. We first estimated how human experts perceive surgery success. Subsequently, we developed a data-driven model integrating these modalities to predict surgery outcomes. The model performance was compared to the current clinical gold standard (three independent human experts) and published outcome calculators. Finally, modality-level phenotypes were derived based on the model’s predictions.

**Results:** Predictions by human experts correlated poorly with post-surgical outcomes, and published outcome calculators did not perform better than the experts (DeLong’s *p* = 0.367). Our model incorporating multimodal data achieved an area under the receiver operating characteristic curve (AUROC) of 0.801. It performed statistically better than the best human expert (DeLong’s *p* = 0.043) and achieved a higher AUROC than the best published surgical outcome calculator (0.801 vs. 0.694).

**Conclusions:** We demonstrated the proof-of-concept that data-driven multimodal phenotypes can inform personalized surgery planning in epilepsy. Furthermore, we provide a framework for integrating multimodal data and benchmarking medical AI performance against human experts.

## Introduction

Epilepsy surgery remains the primary curative intervention for patients with drug-resistant focal epilepsy^1–3^. According to the statistics, only ∼60% of patients achieve seizure freedom^4,5^. In clinical practice, presurgical evaluation involves the collection of multimodal data^6,7^, often including invasive EEG monitoring^8^. Multidisciplinary clinical teams evaluate these data during seizure conferences to formulate an individualized surgery plan. Although experts propose an expected success rate based on statistics, a computational tool for personalized outcome prediction is yet to be formulated. Furthermore, it is unclear how human experts conceive the surgery success or which heuristics they prioritize within this multimodal data. Addressing this challenge is essential, as epilepsy surgery is resource-intensive^7^, carries non-trivial risks, including potential functional deficits^9^, making it critical to distinguish between candidates for surgery and those better suited for alternative therapies such as neuromodulation^10^. This challenge is further complicated by the substantial variation in outcomes, even among patients with apparently the same epilepsy type^5^.

Prior studies aimed at predicting surgery outcomes, have predominantly focused on isolated features, features derived from a single modality^11–15^, or predefined clinical features^16–20^. However, this approach fails to reflect clinical reality; presurgical planning is inherently multimodal, integrating semiology, scalp EEG, neuroimaging, intracranial EEG monitoring, and neuropsychological evaluation. To date, five outcome calculators utilizing predefined clinical features have been proposed in the literature^16^: epilepsy surgery nomogram (ESN, the original version^17^, and the modified version^19^ which included electroencephalographic data), seizure freedom score (SFS)^18^, modified-SFS, and the epilepsy surgery grading scale (ESGS)^20^. The performance of these tools is reasonable^16^; however, these calculators utilize only a limited selection of modalities and features^21^. While these were validated at the population-level, their applicability to specific epilepsy types remains unknown. In addition, these tools remain simplistic, limiting the integration of data from multiple modalities^21^, and may not yet provide the granular, personalized prediction required for patients. Furthermore, previous attempts to integrate multimodal information have demonstrated contradicting results^22,23^, thereby leaving individual risk stratification for epilepsy surgery, a challenge.

We hypothesized that hierarchical integration of multimodal clinical data can predict outcomes non-inferior to or better than the current clinical standard, the human experts. The evaluation was performed using a mesial temporal lobe epilepsy (MTLE) cohort derived from six tertiary epilepsy surgery centers. MTLE is considered a homogenous group associated with the highest surgery success rates, achieving seizure freedom in ∼60-70%^5^ of patients at least 1 year follow-up. Our assumption is that demonstrating an improvement in this ‘best-case’ cohort could imply that such a model can overcome the established performance ceiling of current clinical assessments. This suggests that the model would provide at least a non-inferior benefit for the wider, heterogeneous population of drug-resistant focal epilepsy, where prognostic uncertainty is higher^5^. Our specific objectives were (1) to evaluate how human experts predict surgery success, specifically their inter-rater consistency, and performance in comparison with post-surgery outcomes; (2) to evaluate whether individual features/modalities or existing epilepsy surgery outcome calculators are reliable predictors of outcome; and (3) to develop and validate a model that integrates multimodal data for post-surgical prognosis

## Methods

### Patient cohort and data selection

We utilized a retrospective cohort of 57 unilateral MTLE patients from six centers (in alphabetical order): Centre Hospitalier Universitaire Grenoble-Alpes (France), Duke University Medical Center (USA), Montreal Neurological Institute and Hospital (Canada), St. Anne’s University Hospital (Czech Republic), University Hospital of Nancy (France), and University of Pittsburgh Medical Center (USA). The inclusion criteria were: (1) diagnosis of drug-resistant unilateral MTLE; (2) pre-surgical evaluation using stereo-EEG (SEEG), where SEEG was concordant with a mesiotemporal lobe hypothesis^24^; and (3) subsequent resective or ablative surgery consisting of either laser interstitial thermal therapy (LITT) targeting mesiotemporal structures, or standard/modified anterior temporal lobe resection including mesiotemporal structures, with a minimum post-surgical follow-up of one year. This study was approved by the Duke Institutional Review Board (primary approval, Pro00115198), with secondary approvals obtained from each of the collaborating centers.

Patient demographics and clinical details were derived from the patient charts. In addition, we extracted from SEEG recordings a representative electro-clinical seizure and a 10-minute interictal segment representative of the patient’s interictal epileptiform anomalies. The interictal segment was chosen at least 2 hours away from seizures. The interictal recordings were sampled at a frequency of at least 1 kHz, whereas seizure recordings for some patients were sampled at 200 Hz. We utilized data recorded at higher sampling frequencies whenever available. All data were analyzed in a bipolar montage between adjacent SEEG contacts, and electrode locations were mapped to anatomical regions using the 84-region Desikan-Killiany atlas^25^ (Supplementary S1).

### Evaluation by experts

The clinical standard for epilepsy surgery is based on expert, multidisciplinary review of multimodal data, including electrophysiology, imaging, and neuropsychological assessments, alongside evaluation of functional risks. This process results in a consensus recommendation regarding whether to proceed with a given intervention and, if so, which approach is most appropriate depending on the patient’s expectations. However, it is unknown how experts estimate the likelihood of success. As a first step, we evaluated the diagnostic accuracy of expert prognostication (index test) against the post-surgery outcome (reference standard). Three experts, board-certified epileptologists trained at three different institutions, independently reviewed the pre-surgical data, which includes the patient demographics, types of seizures expressed by the patient, seizure semiology, SEEG segments of a representative seizure and interictal recordings, the anatomical regions corresponding to each SEEG channel, structural MRI findings detailing the presence of lesions, and the type of surgery performed. They were blinded to the post-operative Engel outcomes. The experts assigned a probability of surgical success on a scale of 0-100%. A pre-specified cut-off of ≥50% was defined as a favorable outcome. The confidence of experts in making these predictions was also recorded on a 10-point Likert scale. This study is reported in accordance with the Standards for Reporting Diagnostic Accuracy (STARD 2015) guidelines^26^.

### Input modalities and features

We selected a subset of features with established prognostic significance from the literature. Ten features from four modalities were utilized: type of seizures (one feature), semiology (three features), interictal SEEG (three features), and MRI findings (three features). The types of seizures were categorized into three: focal to bilateral tonic-clonic (FBTC), focal impaired consciousness (FIC), focal preserved consciousness (FPC) seizure^27^. Three features were extracted from semiology: first symptom, rest of the symptoms, and postictal symptoms^28,29^. We utilized three interictal SEEG features that were independent of absolute electrode coordinates. Two features measured the abnormality distance^12^, based on spike^30^ and spike-gamma^11,31^ rates; higher abnormality distances were associated with poor outcomes^12^. The third feature, the spatial perturbation map (SPM), served as a measure of SEEG implantation quality, where higher values correlated with better outcomes^32^. Three features were derived from MRI findings: presence of lesions, region of interest, and lobe of interest. The features were either quantitative (three interictal SEEG features) or categorical (five features). The feature definitions are given in Supplementary S2.

### Comparison with existing outcome calculators

We compared the performance of our model with five surgery outcome calculators published in the literature: ESN^17^, modified-ESN^19^, SFS^18^, modified-SFS, and the ESGS^20^. The description of each of the calculators is provided in Supplementary S3.

### Multimodal feature model

We employed a predictive model where all features and modalities contributed equally to the final decision. The model was comprised of three hierarchical levels: (i) feature-level classification; (ii) modality-level classification; and (iii) a patient-level score. While feature-level classification required model training, the subsequent aggregation levels utilized fixed, parameter-free rules (unweighted averaging) defined a priori. We chose a limited approach because it easily adapts to missing modalities and is appropriate for small datasets. An illustration of the model is given in Figure 1. Further details regarding the model, hyperparameters etc. are provided in Supplementary S4.

**Figure 1:**
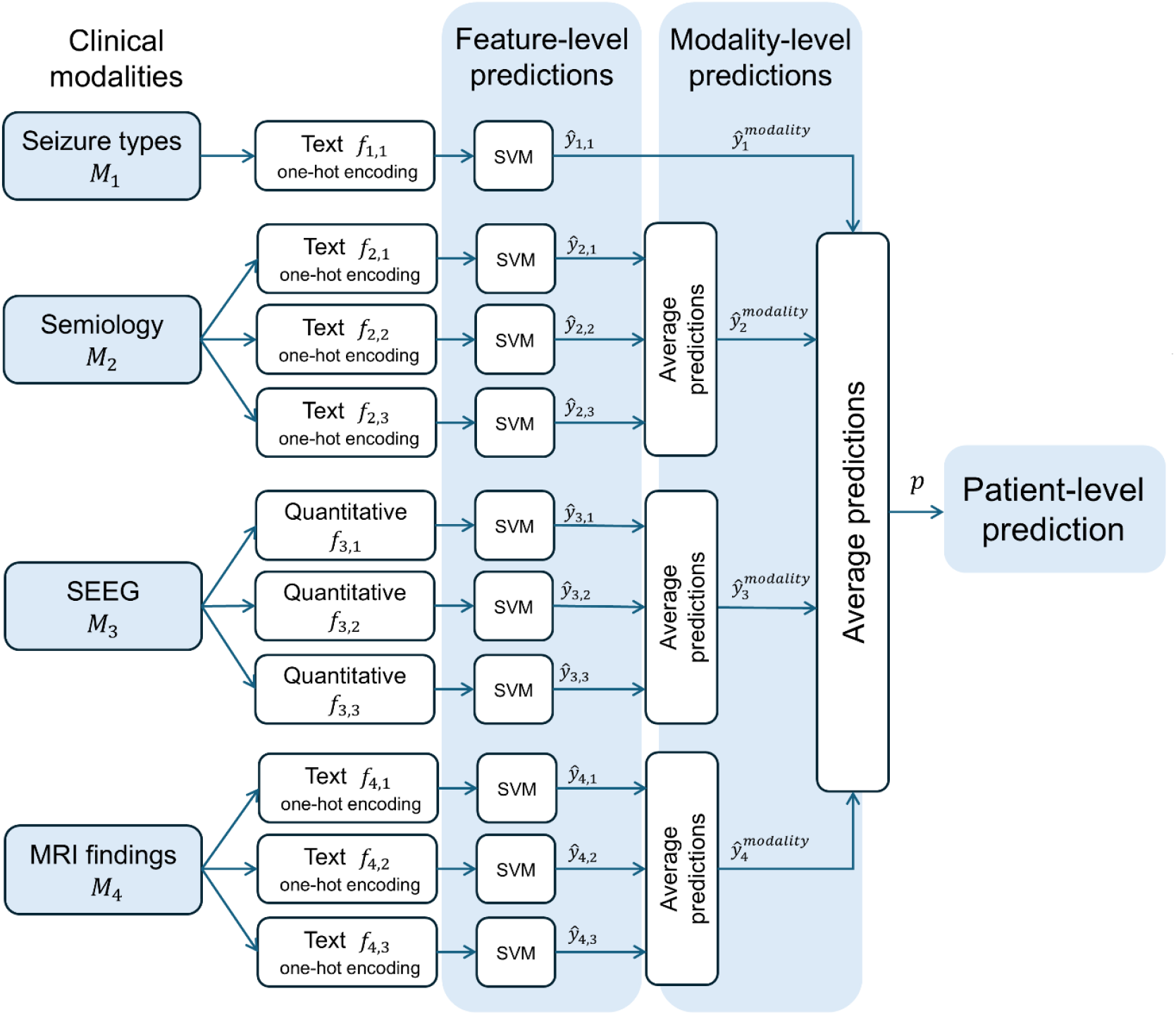
Schematic of the proposed hierarchical model. The model integrates four clinical modalities: seizure types (*M*_1_), semiology (*M*_2_), interictal SEEG (*M*_*3*_), and MRI findings (*M*_4_). All features, except for interictal SEEG, were categorical textual features and were converted into numerical vectors using one-hot encoding. In total, ten features were extracted. The model consists of three levels: (i) feature-level classification utilizing a linear SVM (ŷ _*i,j*_, *i* ∈ {1, 2, 3, 4,} *j* ∈ {1, 2, 3}; (ii) modality-level classification obtained by averaging the predictions of the respective feature SVMs (ŷ_*i*_^*modality*^, *i* ∈ {1, 2, 3, 4}); and (iii) a patient-level prediction derived by averaging the modality-level predictions 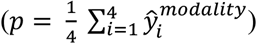. Unlike the feature-level classification, which required training, the subsequent aggregation steps relied on unweighted averaging. A detailed description of the model is provided in Supplementary S4.

**Figure 2:**
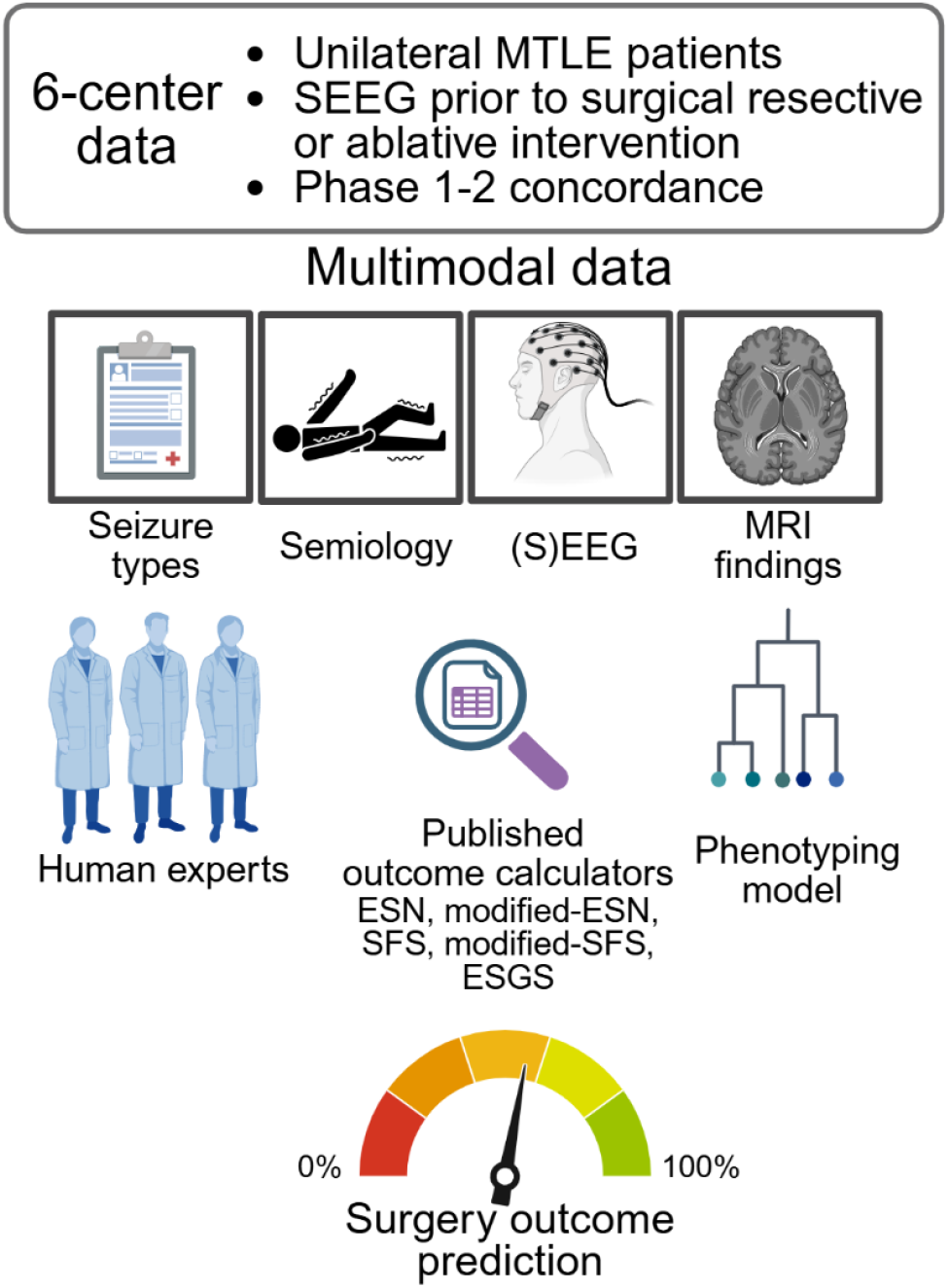
Overview of the study pipeline. The study cohort included patients with unilateral mesial temporal lobe epilepsy from six centers who underwent SEEG investigation prior to resective surgery or ablative intervention. Patients were assessed for Phase 1-2 concordance, defined as the anatomical agreement between non-invasive presurgical evaluations (Phase 1) and invasive SEEG findings (Phase 2)^24^. Using this multimodal dataset, we conducted three evaluations: (a) visual assessment by three board-certified epileptologists trained at different centers; (b) quantitative evaluation using published surgery outcome calculators^1, 17, 18 15^; and (c) a predictive model based on hierarchical integration of four modalities. The three evaluations were assessed against post-surgical Engel classification scores obtained at a minimum follow-up of one year. Created in BioRender. Frauscher, B. (2026) https://BioRender.com/bbpflh5.

### Approach

First, we assessed pairwise inter-rater agreement among experts and evaluated the concordance between their predictions and surgery outcomes. Surgery outcomes were dichotomized into favorable (Engel class I), and non-favorable (Engel classes II-IV) outcomes. Similarly, expert predictions were grouped into favorable (≥50% probability of surgical success) and non-favorable outcomes. These predictions served as the clinical benchmark.

Next, we evaluated the performance of our model by applying leave-one-subject-out (LOSO) cross-validation. The data from all six centers were first pooled together (total *N* = 57). In each trial of cross-validation, we kept the data from a single patient as the left-out test set. Using the remaining *N* − 1 patients as the training data, we implemented the model (Supplementary S4). This model was then evaluated on the left-out test patient. This procedure was repeated for *N* times, and the predictions were aggregated to compute a single receiver operating characteristic curve (ROC) and precision-recall curve (PRC). While LOSO induces bias in many machine learning scenarios when you choose the best performing model of many, here we use default parameters and a single run per split; we therefore have an unbiased estimate of performance and can use standard statistical tests^33^. The statistical tests and performance measures used for analysis are described in Supplementary S5. These performance measures were compared to the clinical standard (three human experts) and the five outcome calculators.

Finally, we defined distinct ‘patient phenotypes’ based on the specific combination of binary predictions generated by the modality-level predictions. We used modality-level rather than feature-level predictions due to sample size constraints. For each resulting phenotype, we calculated the empirical probability of surgery success based on the observed outcomes within our cohort. To visualize how these phenotypes cluster, we generated t-distributed stochastic neighbor embedding (t-SNE)^34^ plots were generated using modality-level predictions as input. The resulting estimated surgery success was then visualized across three categories: high chance (>80%), moderate chance (30-80%), and low chance (<30%).

## Results

### Patient characteristics

The study cohort comprised 57 patients (23 males (40.4%); mean age 32.4 years, range 16-56 years) recruited from six centers: Centre Hospitalier Universitaire Grenoble-Alpes (6), Duke University Medical Center (14), Montreal Neurological Institute and Hospital (4), St. Anne’s University Hospital (10), University Hospital of Nancy (13), and University of Pittsburgh Medical Center (10). Forty patients (70.2%) achieved a favorable outcome (Engel class I). The surgical procedures included anterior temporal lobectomy (39, 68.4%), laser interstitial thermal therapy (10, 17.5%), selective amygdalohippocampectomy (6, 10.5%), and selective temporal pole and amygdala resection (2, 3.5%). Preoperative MRIs were classified as mesial temporal sclerosis (MTS) in 21 (36.8%) patients. Additional bar graphs detailing patient characteristics are available in Supplementary S6.

### Interrater agreement of experts

The experts were confident in their predictions: expert 1 mean 7.5 (range 5-9, 10-point Likert scale), expert 2 mean 8.3 (range 5-9.5), and expert 3 mean 7.2 (range 6-9). Expert 1 predicted a favorable outcome for 50/57 patients, expert 2 for 51 patients, and expert 3 for 54 patients. The mean percentage agreement and Gwet’s AC1 were 93% and 0.57. When evaluating expert predictions against surgery outcomes, we observed a mean percentage agreement of 66.7%, with a Gwet’s AC1 of 0.06 (Table 1). We found no statistical evidence that any of the experts could distinguish between patients with favorable and non-favorable outcomes (p>0.05). Expert 2 demonstrated greater concordance with surgery outcomes than the other two experts (Table 1).

**Table 1.** (A) Results for favorable and non-favorable outcome classification. The results are reported in terms of the area under receiver operating characteristic curve (AUROC), area under the precision recall curve (AUPRC), Wilcoxon rank-sum *p*-values, and Cliff’s *d*. The reference value for AUROC is used as 0.500, AUPRC as 0.702 (proportion of good outcome patients), and 0.05 for *p*-values. The significant *p*-values are highlighted with a *. The results for the best expert, existing surgical outcome calculators^17–20^, and the proposed data-driven model are highlighted in bold. ESN: epilepsy surgery nomogram; SFS: seizure freedom score; ESGS: epilepsy surgery grading scale.

| Classification results |  |  |  |  |  |  |
| --- | --- | --- | --- | --- | --- | --- |
| Expert/Measure/<br>Model | AUROC | AUPRC | Specificity<br>at 80%<br>sensitivity | Balanced<br>accuracy | Statistics between<br>predictions of favorable<br>and non-favorable<br>outcomes |  |
| | | | | | $p$-values | $d$-values |
| Human expert 1 | 0.540 | <0.702 | 17.5% | 48.8% | 0.626 | 0.082 |
| **Human expert 2** | **0.619** | **0.745** | **37.5%** | **58.8%** | **0.155** | **0.238** |
| Human expert 3 | 0.592 | <0.702 | 23.0% | 51.5% | 0.592 | 0.090 |
| ESN | 0.527 | 0.710 | 23.8% | 51.9% | 0.753 | 0.054 |
| **Modified-ESN** | **0.694** | **0.761** | **47.2%** | **63.6%** | **0.021\*** | **0.388** |
| SFS | <0.500 | 0.702 | 15.0% | 47.5% | 0.842 | 0.032 |
| Modified-SFS | <0.500 | 0.702 | 15.0% | 47.5% | 0.842 | 0.032 |
| ESGS | 0.511 | <0.702 | 14.6% | 47.3% | 0.901 | 0.022 |
| **Proposed model** | **0.780** | **0.833** | **63.2%** | **71.6%** | **<0.001\*** | **0.560** |

### Performance of surgery outcome calculators

Among the outcome calculators, only the modified-ESN was able to discriminate between favorable and non-favorable outcomes (Table 2). It achieved a medium effect size of 0.388 (*p*=0.021). However, we found no statistical difference between the ROCs of modified-ESN and the predictions of the best expert (expert 2; DeLong’s *p*=0.367).

### Performance of the hierarchical model

Ten features were used to evaluate the model using LOSO cross-validation. None of these individual features were able to discriminate between favorable and non-favorable outcomes (Supplementary S6). The proposed model was able to distinguish favorable and non-favorable outcomes with an AUROC of 0.780, AUPRC of 0.833, and an effect size of 0.560 (Table 2, Figure 3). It performed statistically better than the human experts, with DeLong’s *p*=0.043 (model vs expert 2). The model achieved a higher AUROC (0.780 vs 0.694, Figure 3b), AUPRC (0.833 vs 0.694, Figure 3c), and effect size (0.560 vs 0.388) than the modified-ESN, although this difference did not reach statistical significance (DeLong’s *p*=0.264). In addition, to evaluate performance in terms of sensitivity and specificity, we report the specificity at a fixed sensitivity of 80% (Figure 3b, Table 3). The model demonstrated higher specificity (63.2%) for predicting outcomes than both the modified-ESN (47.2%) and expert 2 (37.5%; Figure 3b) for a sensitivity of 80%. Further details regarding the individual feature SVMs and their feature importance are provided in Supplementary S7.

**Figure 3:**
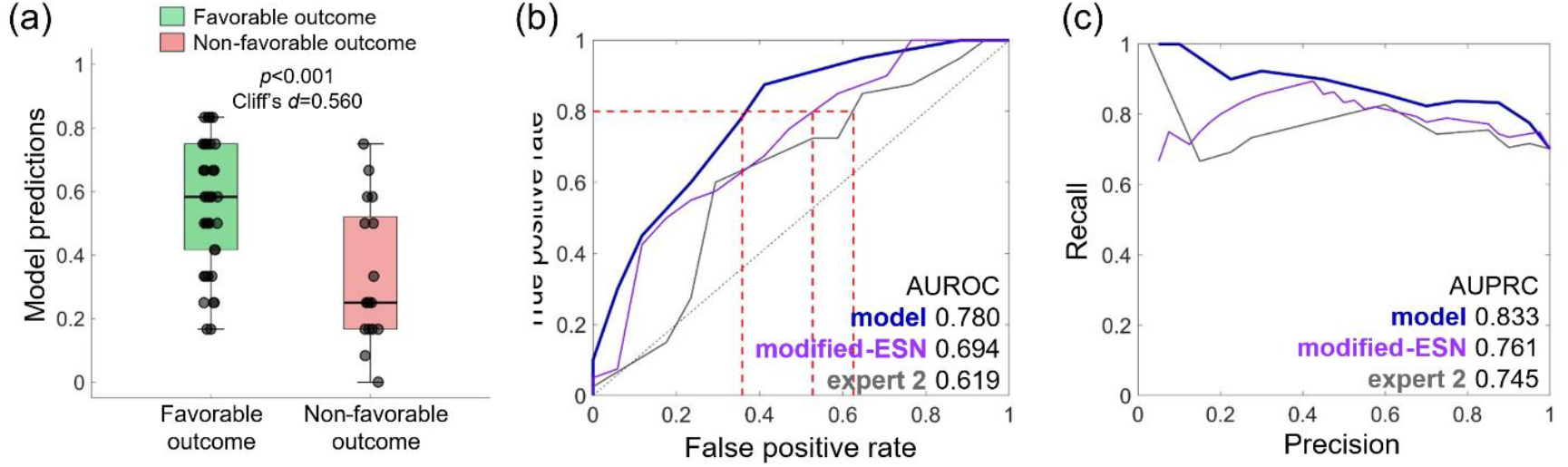
Performance comparison the proposed model to the best expert and outcome calculator: (a) boxplots of predictions of the model for favorable (green) vs. non-favorable (red) outcomes; (b) Receiver operating characteristic curve (ROC) and area under the ROC (AUROC) for the model in comparison with the modified-ESN, and the best expert (expert 2); (c) Precision-recall curve (PRC) and area under PRC (AUPRC) for the model in comparison with the modified-ESN, and the best expert (expert 2). The proposed model was able to discriminate between patients with favorable and non-favorable outcomes (*p*<0.001, Cliff’s d=0.560), outperforming the modified-ESN and expert 2.

### Patient phenotypes

We derived patient phenotypes using the binary predictions from the modality-level classification. Figure 4a illustrates the predictions for each modality, the patient count within each phenotype (subgroup), and the corresponding estimated success rates based on the current dataset. For example, in phenotype 1, all four modalities predicted a favorable outcome. There were three patients with this phenotype, and all three had an Engel 1 outcome, indicating a 100% success rate. The interpretation of the modality-level predictions (i.e., when a modality-level prediction indicates a favorable outcome) is provided in Supplementary S8. For example, in the case of the seizure type modality, a favorable outcome was predicted when a patient had FPC seizures and/or a low frequency of FBTC seizures. Theoretically, four modalities yield 16 (2^4^) possible subgroups; our dataset only contained 15 of these phenotypes. Eight phenotypes were associated with >80% success rate (high), six with a moderate success rate (30-80%), and one with a rate of ≤30% (low). Subsequently, we computed t-SNE plots using these four predictions as inputs. We overlaid the patient groups based on (1) favorable vs. non-favorable outcome (Engel class I vs. II-IV) and (2) the three success rates (high, moderate, and low; see Figure 4b). The t-SNE projection based on these four predictions successfully separated the clusters according to both Engel outcomes (Figure 4b, top) and the estimated success rates (Figure 4b, bottom).

**Figure 4:**
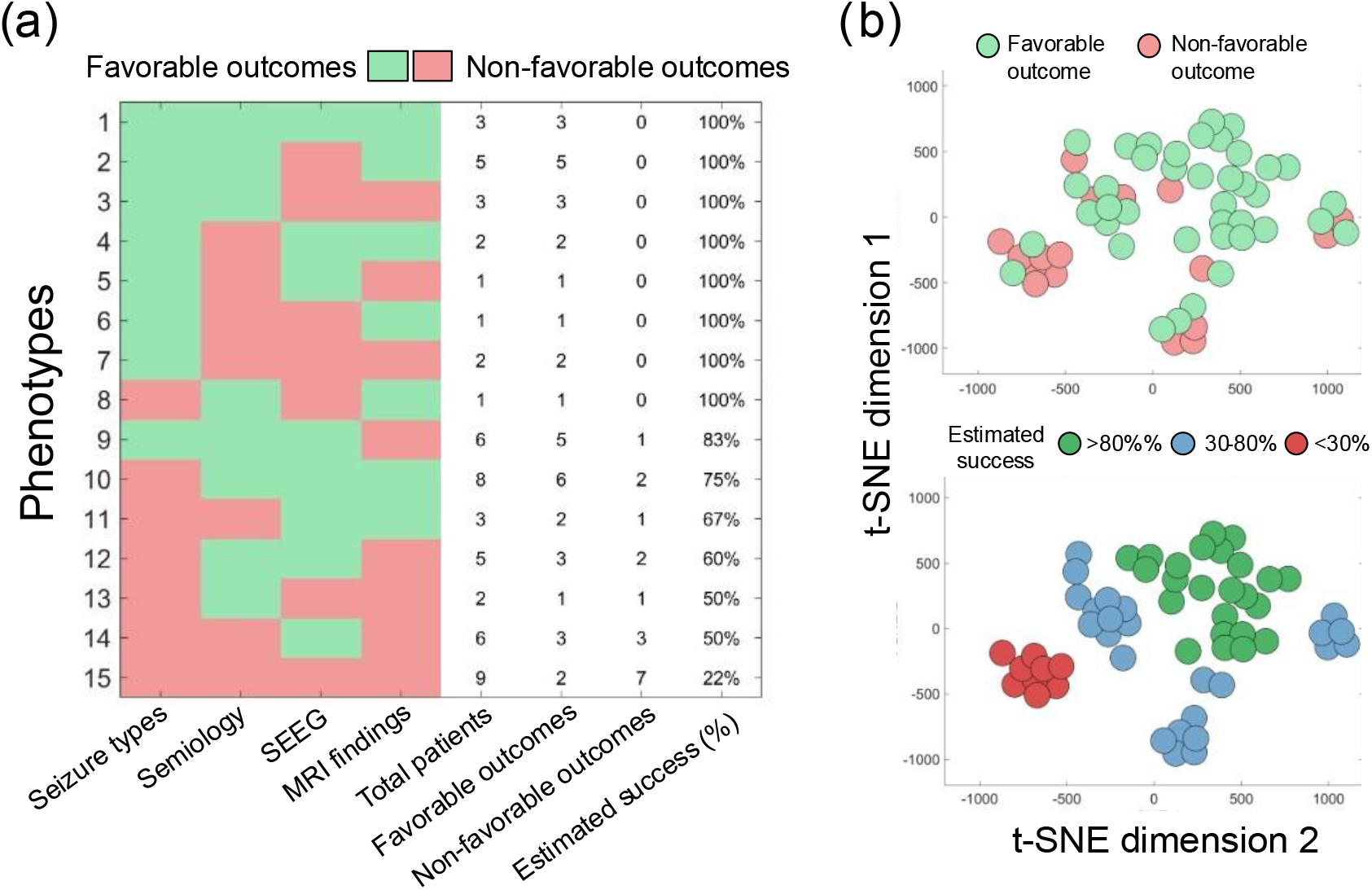
Identification of patient phenotypes and visualization using t-SNE plots: (a) Patient subgroups (phenotypes) derived from the binary predictions of the four modalities. Each row displays the specific combination of modality predictions, i.e., whether a modality predicted that the outcome is going to be favorable (green) or not (red), the number of patients in each subgroup, and the estimated surgical success rate. The interpretation of phenotypes via modality-level predictions is given in Supplementary S8. Phenotypes were categorized into three tiers based on estimated success: high (>80%), moderate (30-80%), and low ≤30%). (b) t-SNE projection of the four modality-level predictions. The two-dimensional representation demonstrates the separation of patient clusters according to surgery outcomes (Engel I vs. II-IV) and the estimated success rate (high, moderate, low).

## Discussion

Prognostication of outcomes in epilepsy surgery remains a challenge. In current clinical practice, prediction relies primarily on visual assessment and the clinical experience of experts. In this study, we demonstrate that hierarchical integration of multimodal features can predict surgery outcomes. The three key findings of this study are: (i) predictions of human experts, the current gold standard, correlated poorly with Engel outcomes; (ii) existing calculators failed to achieve statistically better performance than the experts, and (iii) the hierarchical integration of multimodal features demonstrated a reliable prognostication of epilepsy surgery outcome.

### Implications for clinical practice

Our study is unique in its design, utilizing a specific, epilepsy type-controlled (MTLE), real-world, cohort derived from six tertiary epilepsy surgery centers. We demonstrated that a model incorporating multimodal information performs better than both human experts and published outcome calculators^17–20^. However, it should be noted that while the model showed improved performance, it did not reach statistical significance over the best outcome calculator. This may, in part, reflect the limited sample size and the relatively homogeneous MTLE cohort, which represents a best-case clinical scenario with inherently high success rates^5^. Conversely, this performance gain in the MTLE group could imply that the approach could be effective for the heterogeneous, drug-resistant epilepsy populations. In addition, the varying outcomes among these phenotypes suggest underlying pathophysiological differences within MTLE, which may warrant selective treatment. Ultimately, this work provides a proof-of-concept for hierarchical multimodal integration, demonstrating the potential for data-driven phenotyping to enhance risk stratification and personalized surgery planning.

### Performance of human experts in predicting epilepsy surgery outcomes

We evaluated the prognostic performance of three board-certified epileptologists. Previous research on expert prognostication of epilepsy surgery outcome is limited and contradictory. While one retrospective study of 200 patients reported an accuracy of 83% for a team of experts^23^, a second study of 20 experts (20 patients) reported an expert AUROC of 0.476, indicating performance equivalent to random chance^22^. Both these studies were evaluated on heterogeneous epilepsy populations. In our study, although the experts demonstrated high inter-rater consistency (concordance with each other, AC1=0.57), their predictions correlated poorly with surgery outcomes (AC1=0.06, Table 1). This discrepancy highlights the inherent difficulty of manually synthesizing high-dimensional data, underscoring the need for quantitative^12,13,31^ presurgical assessment of epilepsy.

### Hierarchical integration of multimodal features was able to predict outcomes

The performance of the hierarchical model offers insights into integrating multimodal data. The proposed pipeline provides a framework for handling quantitative and categorical features within small-sample datasets, addressing a vital challenge in medical outcomes research. Interpretability of the data-driven model is essential for clinical adoption. Unlike ‘black box’ algorithms, the proposed structured integration of multimodal information allows epileptologists to understand how the model arrives at predictions, enabling them to validate or challenge specific feature-level and modality-level decisions, based on domain expertise. The feature importance of the feature-level SVM classifiers is detailed in Supplementary S7, and the estimated phenotypes associated with a favorable outcome are shown in Supplementary S8. These interpretations may also advance our understanding of MTLE pathophysiology and introduce a novel framework that can be applied to other epilepsy types. Such transparency is a prerequisite for the deployment of AI in high-stakes clinical decision-making, such as epilepsy surgery.

### Independent features and published calculators failed to predict outcomes

We evaluated three quantitative interictal SEEG features^11,12,31^, a subset of standard clinical modalities, and five established calculators^17–20^. In our cohort, none of these achieved statistically significant AUROC (DeLong’s p>0.05) in comparison to human experts (Table 1, Supplementary S5). A major limitation of these existing biomarkers is that they were derived from a heterogeneous population of drug-resistant epilepsy patients. Since data-driven models optimize for the most predictable outcomes, the derived biomarkers are often disproportionately skewed toward etiologies with inherently higher success rates, such as focal cortical dysplasia^5,11,35^. This study represents the first epilepsy-type (MTLE) controlled study to evaluate these published biomarkers.

### Potential limitations

Our dataset consisted of 57 patients, which limited the statistical power for sub-group analyses and the evaluation of feature-level phenotypes. Utilizing our current findings, we have estimated the sample size necessary to develop a clinically deployable model for future research (Supplementary S9). Second, although comprehensive clinical data was available, we restricted the model to four modalities to mitigate overfitting and reduce dimensionality. While standard clinical evaluations heavily rely on seizure features, we utilized interictal features, as these demonstrated better performance than seizure features in our prior studies^11,12,31,32^. Third, it is important to highlight that the clinicians only had access to selected retrospective segments of ictal and interictal SEEG data and details available from clinical reports. This restricted data access, the heterogeneity in surgical approaches and follow-up durations across centers, and the varied training backgrounds of the experts could potentially lead to deviations from their standard clinical routine and contribute to poor predictions^24^. Finally, we employed simple model using average-based predictions. With larger, sizeable datasets in the future, we aim to learn specific features and modality weights tailored to distinct epilepsy types.

## Conclusion

Epilepsy surgery success rates have stagnated, despite considerable efforts to improve patient care. Our results provide a potential solution for such high-stakes interventions where the dataset is limited: multimodal hierarchical integration of data. Utilizing a diverse, six-center dataset controlled for epilepsy-type (MTLE), we demonstrated the proof-of-concept that data-driven multimodal phenotypes can inform personalized surgery planning. In addition, given approximately the same amount of information and this limited dataset, the AI system performed better than human experts. Consequently, this work represents a crucial step towards personalized phenotype-based treatment for epilepsy surgery.

## Data Availability

All data produced in the present study are available upon reasonable request to the authors.

