## Supplementary material for "Hierarchical integration of multimodal clinical data to predict epilepsy surgery outcome"

**S1. Mapping contacts to brain regions**

The implanted SEEG electrode locations (i.e., three-dimensional coordinates) were derived using preoperative MRI scans and post-implantation CT images. A similar pipeline was followed at the independent centers.

For data from the Centre Hospitalier Universitaire Grenoble-Alpes (France), the Montreal Neurological Institute and Hospital (Canada), and the University Hospital of Nancy (France), coordinates were derived from multisequential and multiplanar MRI or CT data using the MINC toolkit (GNU General Public License)[1]. At Duke University Medical Center (USA), structural T1-weighted MRI scans (with a preference for MPRAGE, SPGR, and MP2RAGE sequences) were processed using FreeSurfer version 7.4.1 for cortical and subcortical segmentation. Pre-implantation MRI and post-implantation CT images were co-registered using BrainLab (version 3.4.2.593). Electrode contacts were then reconstructed in CURRY 8.

All data were analyzed in a bipolar montage referencing adjacent SEEG contacts, and electrode locations were mapped to anatomical regions using the 84-region Desikan-Killiany atlas[2] using built-in Brainstorm routines[3].

**S2. Definition of features**

We selected ten features from four modalities: type of seizures (one feature), semiology (three features), interictal SEEG (three features), and MRI findings (three features). The types of seizures were categorized into three[4]:

- focal to bilateral tonic-clonic (FBTC) seizure
- focal impaired consciousness (FIC) seizure
- focal preserved consciousness (FPC) seizure.

Three features were extracted from semiology[5,6]:

- first symptom
- rest of the symptoms
- postictal symptoms

The semiology was categorized according to the International League Against Epilepsy (ILAE) classification[5], comprising six main categories: autonomic, complex motor, elementary motor, affective (emotional), sensory, and cognitive phenomena. The postictal symptoms included unresponsiveness, language dysfunction, psychiatric signs, headache, and nose wiping.

Three interictal SEEG features that were independent of absolute electrode coordinates were employed:

- abnormality spike distance[7]
- abnormal spike-gamma distance[8]
- spatial perturbation map (SPM)[9]

Spikes and spikes with preceding gamma activity are established biomarkers of epilepsy[7,8,10]. We combined the spike and spike-gamma rates with the newly proposed abnormality distance[11] to account for the focality of the abnormality. Abnormality distance was defined as the weighted standard deviation of the event rates[11]. A higher abnormality distances were associated with poor outcomes[11]. The third feature, the SPM, served as a measure of SEEG implantation quality, where higher values correlated with better outcomes[9].

Three features were derived from MRI findings:

- Presence of lesions:
  - Lesional
  - Abnormal non-lesional
  - Normal
  - Uncertain lesion
- Region of interest:
  - Mesiotemporal
  - Mesiotemporal and neocortical
  - Neocortical
  - Unspecific/Normal
- Lobe of interest:
  - Temporal
  - Extratemporal
  - Temporal and extratemporal
  - Unspecific/Normal

### S3. Surgery outcome calculators

Five surgery outcome calculators were evaluated in this study[12]:

- epilepsy surgery nomogram (ESN)[13]
- modified-ESN[14]
- seizure freedom score (SFS)[15]
- modified-SFS
- epilepsy surgery grading scale (ESGS)[16]

#### S3.1 Epilepsy surgery nomogram

The Epilepsy Surgery Nomogram (ESN)[13] consists of the following features: preoperative seizure frequency (per month, range 0-300), the presence of generalized tonic-clonic (GTC) seizures (yes/no), cause of seizures (mesial temporal sclerosis, malformations of cortical development, stroke, tumor, other), type of surgery (temporal lobectomy, frontal lobectomy, posterior quadrant resection), epilepsy duration at the time of surgery (in years, range 0-65), and gender (male/female). The second version, the modified-ESN[14], includes three additional features: MRI findings (abnormal, normal), EEG seizure localization (sometimes non-localizable, always localizable), and interictal epileptiform discharges (>80% unilateral, bilateral, no epileptiform discharges). In our study, we specifically utilized the '2-Year Probability for an Engel Score' from the ESN and the '2-Year Probability of Seizure Freedom' from the modified-ESN; for both metrics, a higher value indicates a greater likelihood of a favorable surgery outcome. The outcome calculator is available at https://riskcalc.org/FreedomFromSeizureRecurrenceAfterSurgery/.

S3.2 Seizure freedom score
The Seizure Freedom Score (SFS)[15] is calculated using four binary clinical features, where the presence of the feature ('Yes') assigns a score of 0, and its absence ('No') assigns a score of 1. These features include: a normal MRI, > 20 seizures per month, an epilepsy duration of>5 years, and a history of generalized tonic-clonic (GTC) seizures. The modified SFS (m-SFS)[15] builds upon SFS by incorporating two additional features utilizing the same scoring system (Yes = 0, No = 1): the use of invasive evaluation and extratemporal lobe involvement. A total score is then derived by summing the points across the respective variables. A lower score indicated a favorable outcome.

S3.3 Epilepsy surgery grading scale
The Epilepsy Surgery Grading Scale (ESGS)[16] calculates a prognostic score by evaluating five clinical categories. Points are assigned as follows: IQ (<70 yields -1 point; ≥70 or unknown yields 0); semiology (unilateral focal motor activity yields -3; other or unknown yields 0); MRI findings (unilateral MTS yields 5; other temporal lesion yields 3; normal or extratemporal lesion yields 2; ≥2 potentially epileptogenic lesions yields 1); EEG findings (unifocal temporal yields 3; normal, unifocal extratemporal, or bilateral independent/multifocal yields 2; bisynchronous or generalized yields 1); and concordance (concordant yields 2; partially concordant yields 1; not concordant yields 0). A total score is derived by summing the points across these five categories, with a higher total score indicating a greater likelihood of a favorable surgical outcome.

**S4. Multimodal feature model**
We employed a simple predictive model where all features and modalities contributed equally to the final decision. The model comprised three hierarchical levels: (i) feature-level classification; (ii) modality-level classification; and (iii) a patient-level score. While feature-level classification required model training, the subsequent aggregation levels utilized fixed, parameter-free rules (majority voting and unweighted averaging) defined a priori. Let $\boldsymbol{M} = \{M_1, M_2, M_3, M_4\}$ denote the set of four modalities: seizure types, semiology, interictal SEEG, and MRI findings. For each modality, we extracted a feature set $F_{M_i} = \{f_{i,1}, f_{i,2}, \dots, f_{i|F_i|}\}$ for $i \in \{1,2,3,4\}$. In total, ten features were extracted. The features in modalities, seizure types, semiology, and MRI findings were categorical: $f_{i,j} \in C_{i,j} = \{c_1, c_2, \dots, c_{k_{i,j}}\}$ where $k_{i,j} \geq 2$ represented the number of distinct categories for that feature. The interictal SEEG features were numerical: $f_{i,j} \in \mathbb{R}$. The categorical features were transformed into numerical vectors by applying one-hot encoding. The transformation is applied, $\varphi_{i,j}(f_{i,j}): C_{i,j} \rightarrow \{0,1\}^{k_{i,j}}$ such that

$$\varphi_{i,j}(f_{i,j}) = [I(f_{i,j} = c_1), \dots, I(f_{i,j} = c_{k_{i,j}})],$$

where $I(.)$ is the indicator function,

$$I(f_{i,j} = c_p) = \begin{cases} 1 & if\ f_{i.j} = c_p \\ 0 & otherwise \end{cases} .$$

Then, we developed a three-level hierarchical ensemble model to integrate predictions from a feature-level granularity to a global patient-level decision.

*Feature-level classification:* For every feature $f_{i,j}$, we trained an independent binary support vector machine (SVM) classifier, denoted as $h_{i,j}(.)$. A linear kernel SVM with default parameters, with feature standardization, was applied. Given the class imbalance, we employed random under-sampling to ensure a balanced dataset, i.e., an equal number of observations per class.

$$\hat{y}_{i,j} = h_{i,j}\left(f_{i,j}\right) \in \{0,1\} \quad \text{for } j \in \{1,2,3\},$$

where $\hat{y}_{i,j}$ is a prediction for each feature in the dataset.

*Modality-level classification:* If a modality contained more than one feature ($|F_i| > 1$), we applied average voting across the feature predictions:

$$\hat{y}_i^{modality} = \begin{cases} \frac{1}{|F_i|}\sum_{j=1}^{|F_i|} \hat{y}_{i,j} & if\ |F_i| > 1 \\ \hat{y}_{i,j} & otherwise \end{cases},$$

Conversely, for modalities comprising a single feature, the output was that feature's prediction: $\hat{y}_i^{modality} = \hat{y}_{i,j}$.

*Patient-level score:* The final patient-level prediction $p$ averaged the predictive values across all four modalities:

$$p = \frac{1}{4}\sum_{i=1}^{4} \hat{y}_i^{modality}$$

### S5. Statistical and performance measures

The pairwise interrater agreement was evaluated by calculating the percentage agreement ($P_a$) and chance-adjusted Gwet's AC1.[17,18] AC1 was calculated as $(P_a - P_c)/(1 - P_c)$, where $P_c$ is the chance agreement. We utilized standard convention for interpreting AC1; 0-0.20 (slight), 0.21-0.40 (fair), 0.41-0.60 (moderate), 0.61-0.80 (substantial), and 0.81-1.00 (almost perfect).[19] The normality of feature distributions was evaluated using the Kolmogorov-Smirnov test[20]. As the data were not normally distributed, we applied the non-parametric Wilcoxon rank-sum to compute the *p*-values, and Cliff's delta[21] to compute the effect sizes. We followed established guidelines for the interpretation of Cliff's delta[22]: negligible (<0.15), small [0.15-0.33), medium [0.33-0.47), large (>0.47). Classification results were presented in terms of the receiver operating characteristic curve (ROC), area under ROC (AUROC), precision-recall curve (PRC), area under PRC (AUCPRC), sensitivity, specificity, and balanced accuracy. We applied DeLong's test[23] to compare the AUROC across different features/feature combinations. In addition, we report the performance in terms of specificity, and balanced accuracy for a threshold of 80% sensitivity, a widely adopted threshold in medical AI, providing an estimate of model performance at a high-sensitivity operating point[24,25].

### S6. Analysis of individual features

None of the four modalities or ten features individually achieved statistical significance in discriminating between outcome groups. However, distinct trends were apparent across these features. FPC seizures were observed more frequently in patients with a favorable outcome compared to patients with non-favorable outcomes (16/40 vs 3/17, Figure S1). Regarding MRI findings, patients presenting with neocortical abnormalities predominantly achieved favorable outcomes (7/40 vs 0/17, Figure S2b). This could potentially indicate that the abnormalities were identified in the MRI and removed during the intervention. In semiology, cognitive and sensory phenomena were observed in patients with a favorable outcome (6/40 vs 0/17, and 7/40 vs 1/17, Figure S3a), whereas complex motor phenomena were identified exclusively in patients with a non-favorable outcome (0/40 vs 2/17, Figure S3a). Postictal language dysfunction was also more common in patients with a favorable outcome (18/40 vs 3/17, Figure S3c). None of the quantitative interictal SEEG features statistically discriminated between outcome groups: spike abnormality distance ($p$=0.206), gamma abnormality distance ($p$=0.166), and SPM ($p$=0.547), Figure S4.

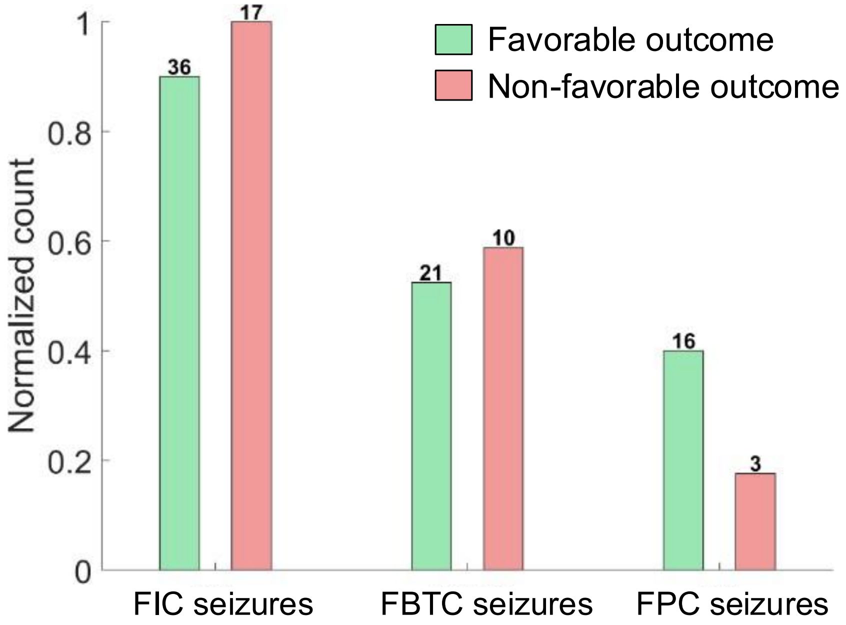


Figure S1: Bar graphs showing the proportion of seizure types across patients with favorable and non-favorable surgery outcomes. The favorable outcomes are shown in green and the non-favorable outcomes in red. The bars are sorted in descending order of prevalence. The y-axis represents the normalized count (proportion) of patients, with values displayed above each bar. FIC: focal impaired consciousness; FBTC: focal to bilateral tonic-clonic; FPC: focal preserved consciousness.

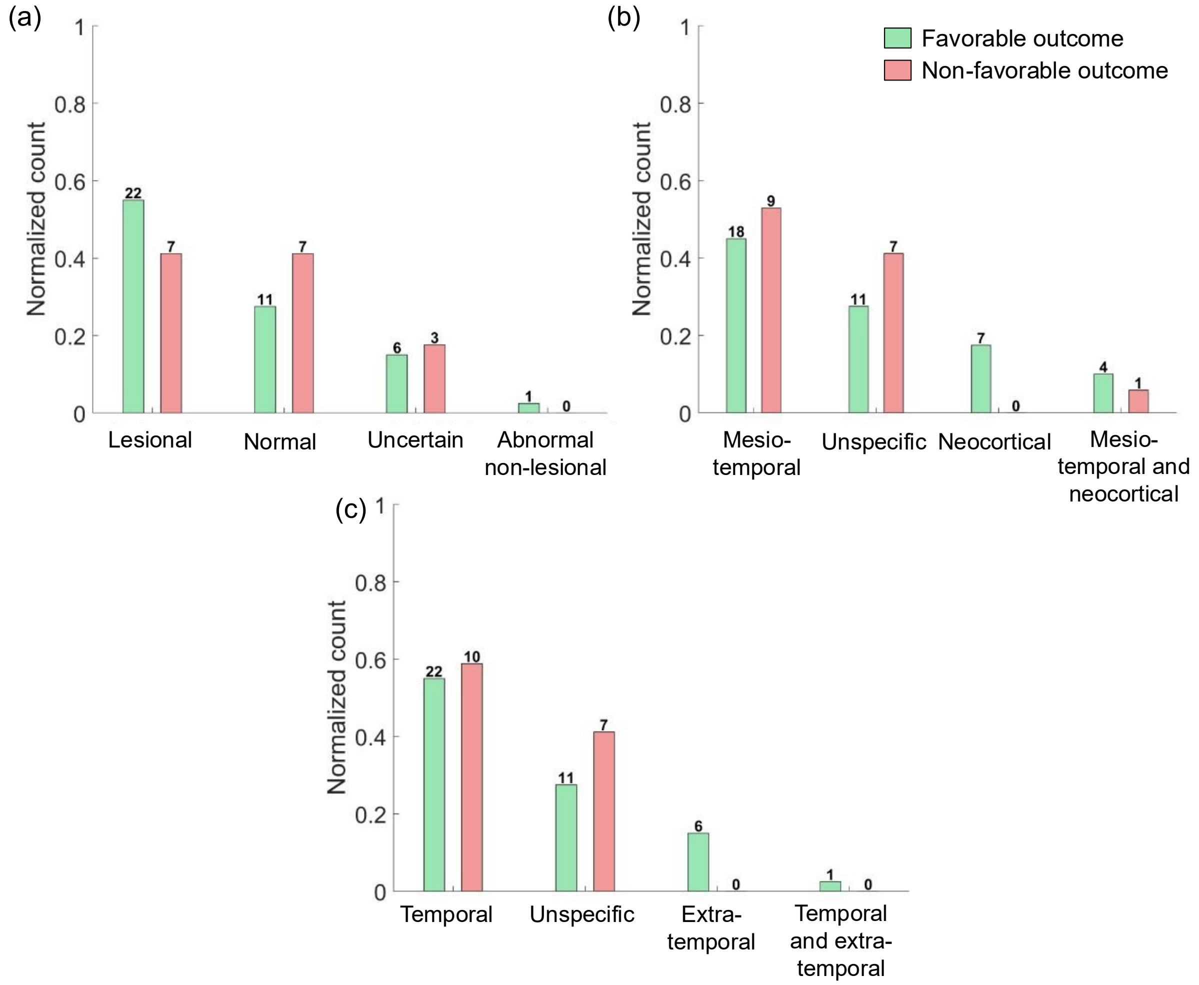


Figure S2: Bar graphs showing the proportion of Magnetic resonance imaging (MRI) findings: (a) presence of lesions, (b) region of interest, and (c) lobe of interest, across patients with favorable and non-favorable surgery outcomes. The favorable outcomes are shown in green and the non-favorable outcomes in red. The bars are sorted in descending order of prevalence. The y-axis represents the normalized count (proportion) of patients, with values displayed above each bar.

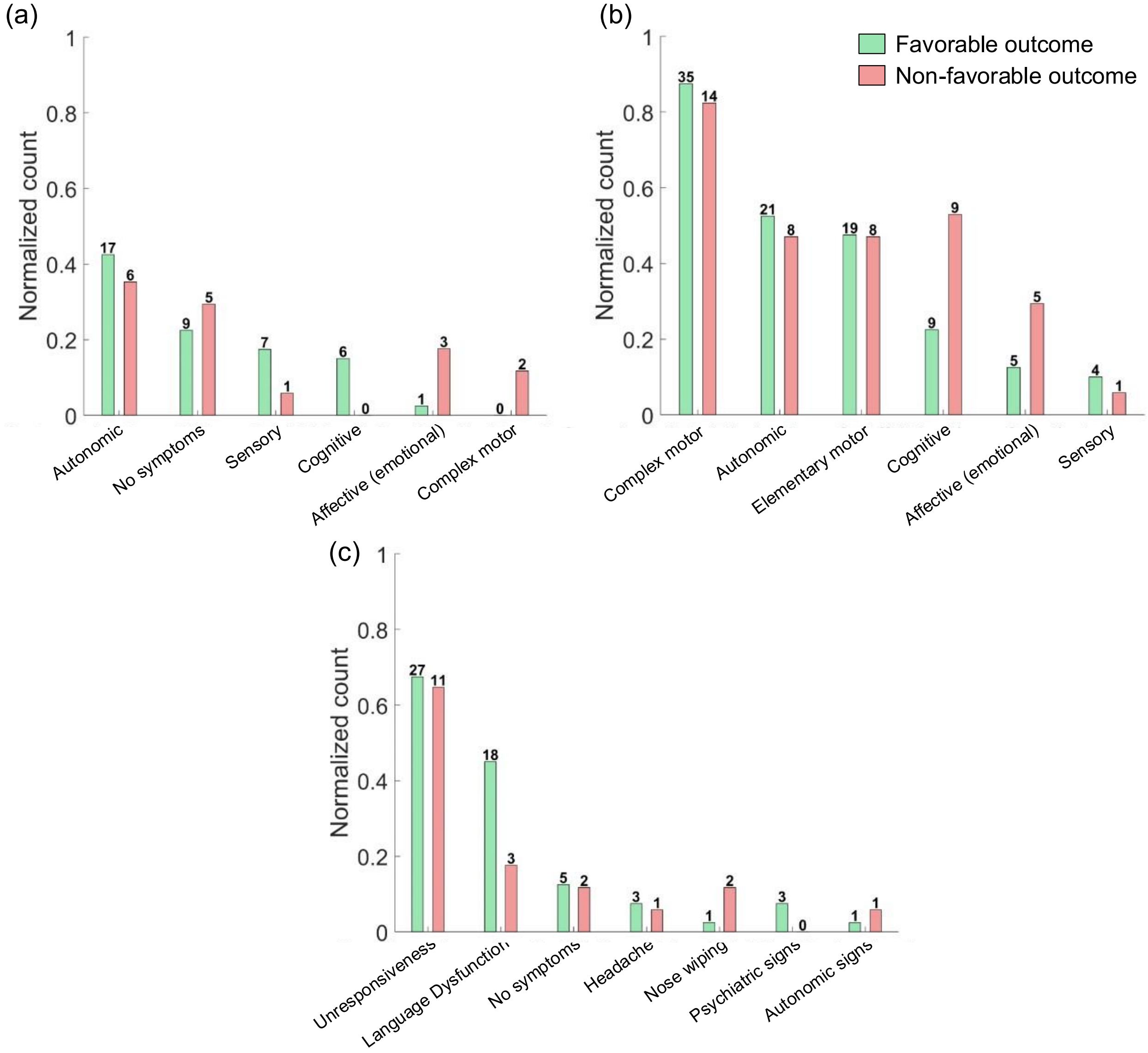


Figure S3: Bar graphs showing the prevalence of semiology (a) first symptom, (b) rest of symptoms, and (c) postictal symptoms, across patients with favorable and non-favorable surgery outcomes. The favorable outcomes are shown in green and the non-favorable outcomes in red. The bars are sorted in descending order of prevalence. The y-axis represents the normalized count (proportion) of patients, with values displayed above each bar.

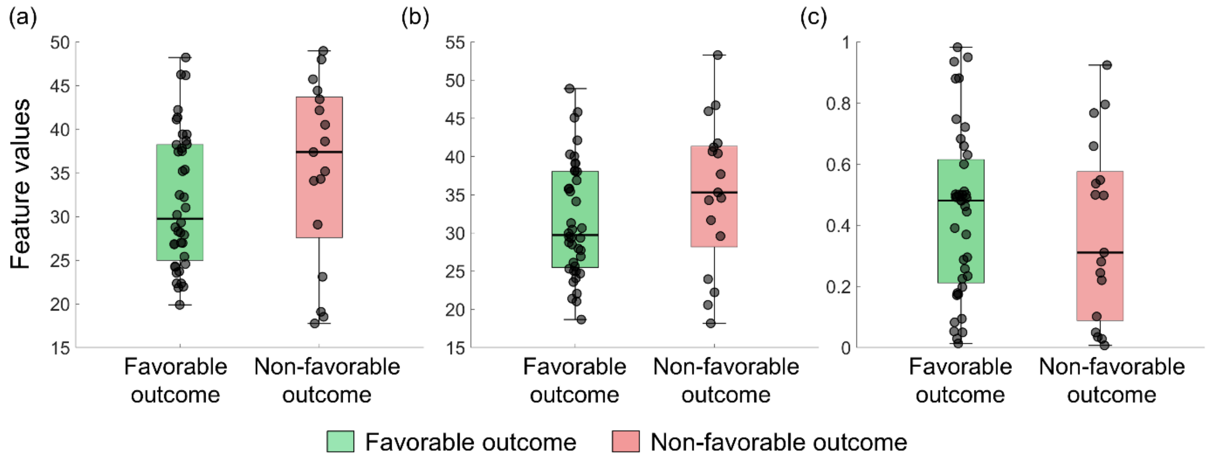


Figure S4: Boxplots for Stereo-EEG interictal features: (a) abnormality spike distance, (b) abnormality spike-gamma distance, and (c) spatial perturbation map. The favorable outcomes are shown in green and the non-favorable outcomes in red. None of the features were able to statistically discriminate the outcome groups: spike abnormality distance (p=0.206), gamma abnormality distance (p=0.166), and spatial perturbation map (SPM) (p=0.547).

**S7. Feature importance of feature-level classification**

We employed a predictive model where all features and modalities contributed equally to the final decision. In the first level, the feature-level classification, we employed a linear support vector machine (SVM) for each individual feature. We estimated the feature importance (ranking) for both the text and quantitative features during the leave-one-subject-out (LOSO) cross-validation. Text features were converted into numerical arrays using one-hot encoding. The normalized (normalized to 1 in each iteration) values of the SVM weight vector ($\beta$) were used as the importance. For quantitative features, since we implemented an SVM for each feature, the SVM identifies an optimal threshold that separates the favorable and non-favorable outcomes. The importance/threshold was computed for each iteration ($N = 57$ times), and the values were averaged.

In seizure types, the classifier assigned the highest weight to FPC seizures, followed by FBTC seizures (Figure S5). For MRI findings, the highest importance for the presence of lesions feature was for normal and lesional findings (Figure S6a); for the region of interest, there was almost equal importance for mesial-temporal and neocortical, unspecific, and neocortical findings (Figure S6b); for the lobe of interest, it was highest for extra-temporal findings, followed by unspecific, temporal and extra-temporal findings (Figure S6c). In semiology first symptom feature, approximately equal importance was assigned to autonomic phenomena, affective (emotional) phenomena, and the absence of a first symptom (Figure S7a). For the rest of the semiology, cognitive symptoms had the highest importance, followed by affective and sensory phenomena (Figure S7b). For postictal symptoms, language dysfunction had the highest importance, followed by nose wiping (Figure S7c). For quantitative features, the mean thresholds were 35.178 for abnormality spike distance, 36.532 for abnormality spike-gamma distance, and 0.468 for the SPM.

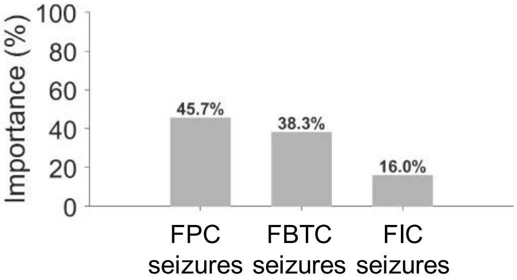


Figure S5: Bar graphs showing the average support vector machine (SVM) feature importance of seizure types. The bars are sorted in descending order of importance. The y-axis represents the feature importance (percentage) with values displayed above each bar. FPC: focal preserved consciousness; FBTC: focal to bilateral tonic-clonic; FIC: focal impaired consciousness.

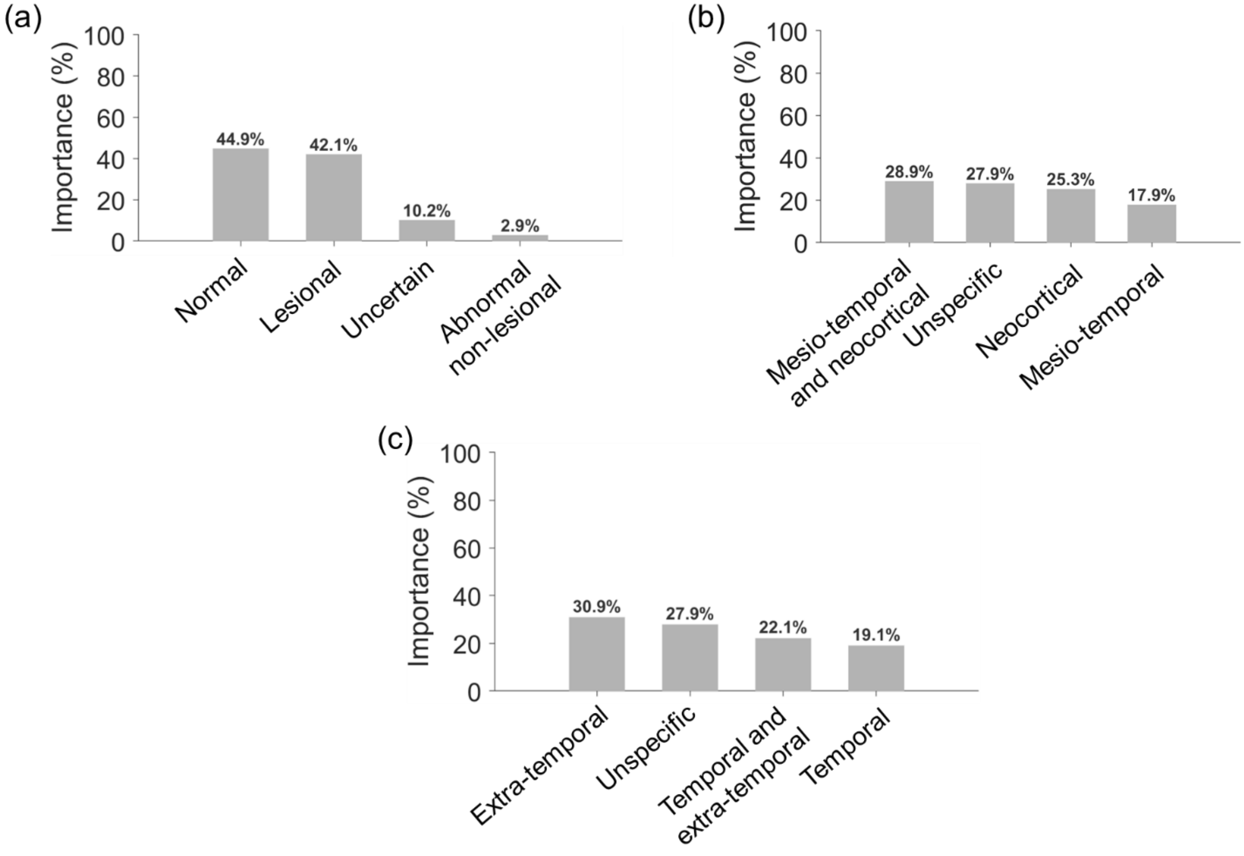


Figure S6: Bar graphs showing the average support vector machine (SVM) feature importance of (a) seizure types and (b) Magnetic resonance imaging (MRI) findings. The bars are sorted in descending order of importance. The y-axis represents the feature importance (percentage) with values displayed above each bar. FPC: focal preserved consciousness; FBTC: focal to bilateral tonic-clonic; FIC: focal impaired consciousness.

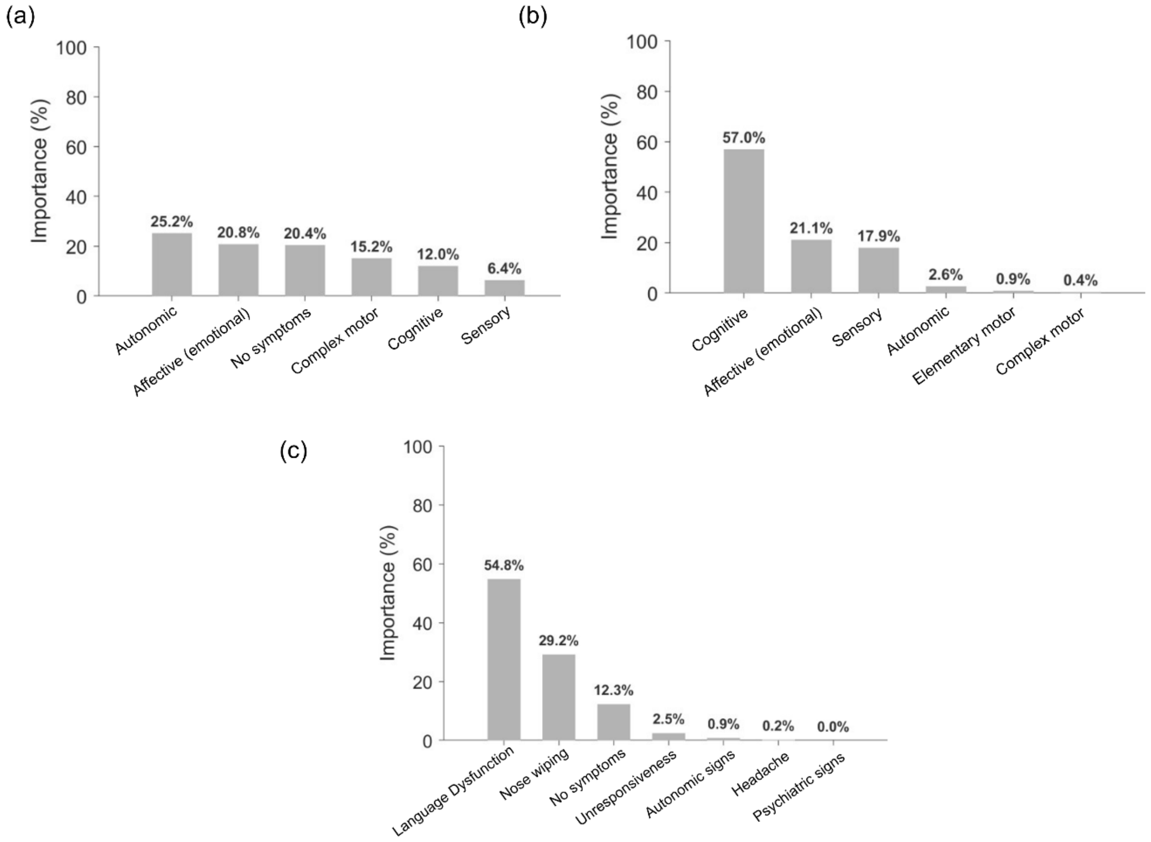


Figure S7: Bar graphs showing the average support vector machine (SVM) feature importance of semiology features: (a) first symptom, (b) rest of symptoms, and (c) postictal symptoms. The bars are sorted in descending order of importance. The y-axis represents the feature importance (percentage) with values displayed above each bar.

**S8. Phenotypes based on modality-level classification**

The patient phenotypes were derived based on modality-level predictions rather than feature-level predictions due to the limited data size. However, using the feature importance for feature-level SVM (Supplementary S6) and analysis of individual features (Supplementary S5), we identified the primary conditions driving a favorable modality-level prediction. These criteria are used to interpret resulting patient phenotypes.

Table S1: Estimated conditions for a modality-level prediction for a favorable outcome.

| **Modality** | **Estimated conditions for the prediction of a favorable outcome** |
|---|---|
| Seizure types | Seizures classified as<br>• exclusively FPC seizures or<br>• a low frequency of FBTC seizures |
| Semiology | At least two of the conditions are satisfied:<br>• first symptom was |

| | |
|---|---|
| | ○ autonomic<br>○ not affective (emotional)<br>○ present<br>○ not complex motor<br>• rest of the symptoms were<br>○ not cognitive<br>○ not affective (emotional)<br>○ sensory<br>• postictal symptoms were<br>○ language dysfunction<br>○ not nose wiping |
| SEEG | At least two of the conditions are satisfied:<br>• Abnormality spike distance <35.178<br>• Abnormality spike-gamma distance<36.532<br>• SPM value>0.468 |
| MRI findings | At least two of the conditions are satisfied:<br>• presence of lesions was<br>○ not normal<br>○ lesional<br>• region of interest was<br>○ neocortical<br>○ mesio-temporal and neocortical<br>○ not unspecific<br>• lobe of interest was<br>○ extra-temporal<br>○ not unspecific<br>○ temporal and extra-temporal |

**S9. Estimating the required cohort for the final model**

To guide the future development of a clinical prediction model, we estimated the required cohort size using closed-form criteria that prioritize minimal overfitting[26–28]. We used a shrinkage factor of 0.9 to limit optimism in predictor effect estimates and required an overall outcome risk within ≤0.05 margin of error[28]. By employing the achieved AUROC of 0.780 and the prevalence of favorable outcomes (0.7 in our cohort), we estimated that 323 patients are necessary to meet performance criteria[28,29]. This calculation utilizes the anticipated model performance to ensure the effective sample size is sufficient, computed via the R pmsampsize package[30]. The calculator is available online at https://riskcalc.org/samplesize/#.
